# Credible learning of hydroxychloroquine and dexamethasone effects on COVID-19 mortality outside of randomized trials

**DOI:** 10.1101/2020.12.06.20244798

**Authors:** Chad Hazlett, David Ami Wulf, Bogdan Pasaniuc, Onyebuchi A. Arah, Kristine M. Erlandson, Brian T. Montague

**Author notes:** KE and BM are co-senior authors.

## Abstract

**Objectives:** To investigate the effectiveness of hydroxychloroquine and dexamethasone on coronavirus disease (COVID-19) mortality using patient data outside of randomized trials.

**Design:** Phenotypes derived from electronic health records were analyzed using the stability-controlled quasi-experiment (SCQE) to provide a range of possible causal effects of hydroxychloroquine and dexamethasone on COVID-19 mortality.

**Setting and participants:** Data from 2,007 COVID-19 positive patients hospitalized at a large university hospital system over the course of 200 days and not enrolled in randomized trials were analyzed using SCQE. For hyrdoxychloroquine, we examine a high-use cohort (n=766, days 1 to 43) and a later, low-use cohort (n=548, days 44 to 82). For dexamethasone, we examine a low-use cohort (n=614, days 44 to 101) and high-use cohort (n=622, days 102 to 200).

**Outcome measure:** 14-day mortality, with a secondary outcome of 28-day mortality.

**Results:** Hydroxycholoroquine could only have been significantly (p<0.05) beneficial if baseline mortality was at least 6.4 percentage points (55%) lower among patients in the later (low-use) than the earlier (high-use) cohort. Hydroxychloroquine instead proves significantly harmful if baseline mortality rose from one cohort to the next by just 0.3 percentage points. Dexamethasone significantly reduced mortality risk if baseline mortality in the later (high-use) cohort (days 102-200) was higher than, the same as, or up to 1.5 percentage points lower than that in the earlier (low-use) cohort (days 44-101). It could only prove significantly harmful if mortality improved from one cohort to the next by 6.8 percentage points due to other causes—an assumption implying an unlikely 84% reduction in mortality due to other causes, leaving an in-hospital mortality rate of just 1.3%.

**Conclusions:** The assumptions required for a beneficial effect of hydroxychloroquine on 14 day mortality are difficult to sustain, while the assumptions required for hydroxychloroquine to be harmful are difficult to reject with confidence. Dexamethasone, by contrast, was beneficial under a wide range of plausible assumptions, and was only harmful if a nearly impossible assumption is met. More broadly, the SCQE reveals what inferences can be credibly supported by evidence from non-randomized uses of experimental therapies, making it a useful tool when randomized trials have not yet produced clear evidence or to provide corroborative evidence from different populations.

## 1 Introduction

Although randomized controlled trials (RCTs) are the gold standard for learning causal effects of treatments on outcomes, running and awaiting the results of RCTs remains challenging and sometimes infeasible. This is particularly evident in the case of the SARS-CoV-2 infection that led to the coronavirus disease (COVID-19) pandemic, where a multitude of treatments were adopted in the clinic on an urgent basis. It is particularly in these cases where the ability to draw credible inferences regarding the effects of treatments used outside of RCTs is of enormous interest to patients, healthcare workers and researchers. Yet, conventional approaches to such observational studies have well-known limitations, particularly their vulnerability to uncontrolled confounding, which can bias results such that harmful treatments could appear beneficial or vice versa without warning. Physicians and other expert consumers of medical research are often (rightly) wary of drawing conclusions about treatment effects – be they null, beneficial, or harmful – from non-randomized comparisons. Nevertheless, particularly in emergencies such as the COVID-19 pandemic, healthcare providers often need to make decisions before RCTs have been completed or for individuals not well represented in those trials. Further, the global response to COVID-19 has seen numerous treatments provided off-label or through emergency access provisions in parallel with ongoing RCTs, raising the question of what can credibly be learned from the experiences of patients receiving these treatments outside of RCTs.

This study employs the stability-controlled quasi-experiment (SCQE)[1, 2] approach to investigate treatment effects on COVID-19 patients, which differs from conventional approaches for observational studies in two key ways. First, unlike standard covariate-adjustment strategies (regression, matching, weighting, and stratification), SCQE does not rely on the assumption that there are no unobserved confounders, i.e. that the treated and untreated groups are comparable after accounting for observed covariates. Instead, SCQE produces estimates that depend only on what the user is willing to assume about the *baseline trend*, here meaning changes in the COVID-19 mortality rates from one cohort to another that are not caused by changes in the treatment in question. Second, whereas conventional approaches present a single estimate and confidence interval that can be correct only under the assumption of no unobserved confounding or other sources of bias, SCQE displays the entire range of estimates obtained over a plausible range of assumptions about this baseline trend. These results can be restated to reveal *what assumptions about the baseline trend in mortality would have to be defended* in order to argue that the treatment was beneficial, null, or harmful. Such an exercise avoids reliance on narrow assumptions. Yet, as illustrated here, it can be informative both in showing the range of plausible effects of a treatment and in showing us what cannot be safely concluded from available evidence.

Using electronic health records from a university hospital cohort of 2,007 patients admitted over 200 days, we apply SCQE to investigate what can be concluded regarding the impact of hydroxychloroquine and dexamethasone on mortality in patients with COVID-19.

## 2 Methods

### 2.1 Approach and assumptions

To build intuition for the SCQE approach, let us consider a “natural experiment” that leverages changes in treatment prevalence over time. Suppose there are two cohorts of patients. In the first, no patients have access to a given treatment, and mortality is 20%. In another cohort (e.g., taken from a later period of time at the same facilities), 50% of patients are administered a new treatment. They do so not at random, but based on patient and physician judgement and choice. Suppose the overall mortality rate in the second cohort is 15%. With an assumption that the two cohorts of patients are comparable (i.e. they would have the same average outcomes, absent treatment differences) we can estimate that being in the second (“high-use”) cohort reduced mortality by 5 percentage points. Further, since all of this benefit comes from the half of patients who opted to take treatment, the benefit per treated patient must be twice that (i.e. a 10 percentage point benefit per treated patient). Note that the required assumption here regards comparability of the cohorts, and not comparability of the treated to the untreated within either the first or the second cohort. This is beneficial as we acknowledge that treatment decisions can be made in part due to unobservable factors, making the treated and control groups incomparable regardless of efforts to adjust for all measured or observed variables.

Such an approach, however, is limited by the assumption that the two cohorts would have the same average mortality rate, absent changes in the treatment. The SCQE takes the more flexible position of allowing the cohorts to differ in this regard by variable, postulated degrees. That is, we allow for some “baseline trend” that describes how differently the cohorts would have fared on their average outcomes, if not for changes in the treatment in question. Equivalently, this baseline trend can be defined as the difference in average outcomes between cohorts that we would have seen if no patients in either cohort had used the treatment. For instance, if treatment changes (other than the one in question) or changes in the composition of the cohorts would have generated a mortality rate that was 2 percentage points lower in the later, high-use cohort than the earlier one, the baseline trend for that analysis would be −2 percentage points.

The key mathematical fact is that in this case, *for any assumed baseline trend, we can estimate the treatment effect experienced by the treated patients*, without additional assumptions or covariates [1, 2]. For intuition behind this result, we return to the natural experiment considered above, where no individual in the first cohort takes the treatment, and hence the average outcome we observe is the “average non-treatment outcome”.^1^ If we add to this the assumed baseline trend, we obtain the average non-treatment outcome in the high-use cohort, i.e. the outcome we would expect if we could see how all individuals in this cohort fared absent the treatment (regardless of whether they actually took the treatment).

Algebraically, this average non-treatment outcome over *everybody* in the second cohort is the sum of two terms: (i) the average non-treatment outcome we observe from the untreated patients in this cohort, times the proportion that were untreated, and (ii) the (unobservable) average non-treatment outcome that the treated patients would have had, times the proportion that were treated. Since the average non-treatment outcome for the treated is the only unknown in this equation, we can solve for this quantity (see [1] for a complete mathematical treatment). Next, the (observed) average treatment outcome for the treated minus this average non-treatment outcome for the treated is the average treatment effect among the treated (ATT). Figure 1 illustrates this reasoning graphically, using values similar to those from the dexamethasone study below.

**Figure 1:**
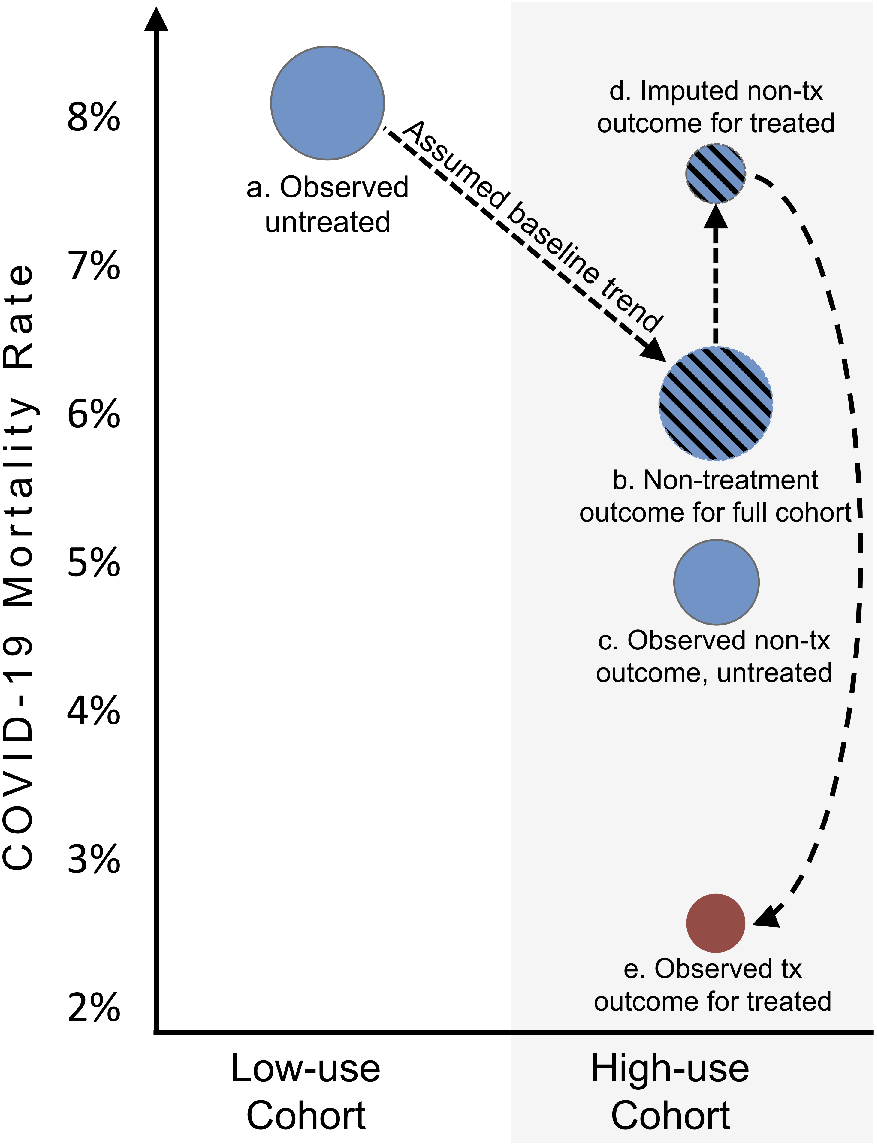
Understanding the SCQE. *Note:* Each ball represents a group, and the height represents that group’s average outcome. Starting on the left, in the low-use (in this case, no-use) cohort we observe the average mortality (8%) under non-treatment. We then impose an assumption regarding how the non-treatment outcome would have changed from one cohort to the next. Here this is a 2 percentage point drop, meaning the average non-treatment outcome over the entire high-use cohort is assumed to be 6% (b). Because the value of (b) is the weighted sum of the average non-treatment outcomes for those who were not treated (c) and those who were treated, we can solve algebraically for the average non-treatment outcome that would have been experienced by the treated (d). Comparing the observed average outcome for the treated (e) to this imputed average non-treatment outcome for the treated (d) produces the average treatment effect for the treated. No assumption regarding the comparability of the treated and control (c and e) is made, only an assumption on the trend in the average non-treatment outcome.

Finally, rather than place our confidence in a single assumption, we “invert” the analysis to reveal *the needed assumptions about the baseline trend in mortality to declare that a given treatment had a beneficial, null, or harmful effect*. Confidence intervals can be constructed for the effect estimate at any given choice of the baseline trend assumption, using the approach described in [2]. Throughout this paper, we describe an estimated effect as a “significantly” beneficial or harmful effect when its 95% confidence interval excludes zero, which is equivalent to a two-sided p-value at or below 0.05.

While no analysis can determine the true value of the baseline trend, beliefs about this quantity can be defended or challenged through auxiliary analyses, such as examining the change in the composition and risk factors of the patients in the two cohorts and changes in any other documented treatment practices. We consider what baseline trends can be deemed plausible or implausible in the Discussion below.

While we have described the approach in its simplest form, several extensions are important, some employed here. First, we need not have a cohort with zero use of the treatment, just two cohorts with sufficiently different levels of treatment.^2^ Second, the two cohorts do not need to be cohorts separated by time; they could be cohorts from separate hospitals, for example. We need only be able to consider how widely the high-use cohort may have differed in its average outcome from the low use cohort, for reasons other than the treatment in question. Third, while we employ individual observations for the analysis and a range of auxiliary variables that are an aid to validating the approach, the SCQE can be used to estimate effects where we are only given average outcomes and the proportion treated in two cohorts.^3^

### 2.2 Data Collection

Data were extracted from the electronic medical records for a multicenter hospital system including an academic tertiary referral hospital. Hospital courses were identified based on a documented COVID-19 infection indicated by either recorded diagnosis or identification of a positive PCR test for the SARS-CoV-2 virus. Data were extracted for all persons with hospitalizations that began between 3/8/2020 and 10/7/2020. Individuals with multiple hospitalizations within the follow-up period were considered to be a single observation, using the earlier date of admission for cohort determination and considering therapies and mortality within the follow-up period if they occurred during any of the hospitalizations.

Clinical data extracted included demographic factors (age, sex, race/ethnicity, body mass index at or prior to the period of hospitalization), baseline laboratory assessments (white blood cell count, C-reactive protein, ferritin, procalcitonin), medication use (remdesivir, convalescent plasma, hydroxychloroquine, dexamethasone, prednisone, methylprednisolone, hydrocortisone), and use of proning for assistance with ventilatory support. We additionally extracted whether the patient had been admitted by transfer from a skilled nursing facility, and disposition at discharge.

Use of dexamethasone and hydroxycholoroquine, our treatments of interest, were defined as any use during the hospital stay(s). Hydroxychloroquine was administered as a standard 5-day course. For dexamethasone, the prescribed course was variable in the few cases given in the low-use cohort (days 44-101). Its prescription in the high-use cohort became more standardized, typically administered at 6 mg for 10 days, following initial evidence for its potential effectiveness at that dose [5].

### 2.3 Cohort Construction

#### Hydroxychloroquine

Initially, hydroxychloroquine was widely used, given to 62% of patients admitted in the first two weeks. Usage then began to fall steadily, with fewer than 2% of patients admitted in week 7 or later receiving it. Cohorts were constructed based on each patient’s day of admission. Data from all days (1 to 200) were first split into two cohorts based on the split-point that would maximize the strength of relationship between cohort and probability of receiving hydroxychloroquine, as judged by the F-statistic. This occurred at day 44. Next, the second cohort was trimmed, to avoid covering a period in which dexamethasone use rose. Ending the second cohort on day 82 minimized the difference in proportion of patients receiving dexamethasone in the first and second cohorts. When used in either cohort, hydroxychloroquine was administered as a standard 5-day course.

#### Dexamethasone

Use of dexamethasone began low and remained at 5% or lower for the first 15 weeks, after which it steadily rose and peaked near 50% in week 21. Cohort construction proceeded by first choosing the split date that maximized the difference in dexamethasone use in the two cohorts, as judged by the F-statistic, which occurred at day 102. We then trimmed the first cohort to begin on day 44, ensuring little change in hydroxychloroquine usage between the cohorts. Dexamethasone use in the first, low-use cohort was largely unstandardized. The transition to higher use in the later cohort was triggered partly by the preliminary report of the RECOVERY trial [5], and followed its standard dosing regimen of 6 mg of dexamethasone once daily for up to 10 days.

We employed the SCQE approach using the `scqe` statistical package for `R` [6].

## 3 Results

### 3.1 Little evidence supporting beneficial role for hydroxychloroquine

The “high-use” (first) cohort for the hydroxychloroquine study included 766 patients admitted from day 1 to 43, of which 36% used hydroxychloroquine, and a “low-use” (second) cohort of 548 patients admitted between days 44 and 82, of which only 2.9% used hydroxychloroquine. The F-statistic for difference in hydroxychloroquine use between cohorts was 242.3 (p<1e-15). Mortality at 14-days was 11.6% in the high-use cohort (89/766) and 8.6% (47/548) in the low-use cohort, for a raw risk difference (RD) of 3.0 percentage points (t=1.79, p=0.07).

Figure 2 shows results for hydroxychloroquine. The vertical axis shows different assumptions regarding the baseline trend, i.e. mortality shifts absent changes in hydroxychloroquine use. These are expressed in terms of the mortality change going from the low-use to high-use cohorts, and because the high-use cohort came first and the low-use came second, a value of 0.02, for example, reflects an improvement (decrease) over time in mortality by 2 percentage points.

**Figure 2:**
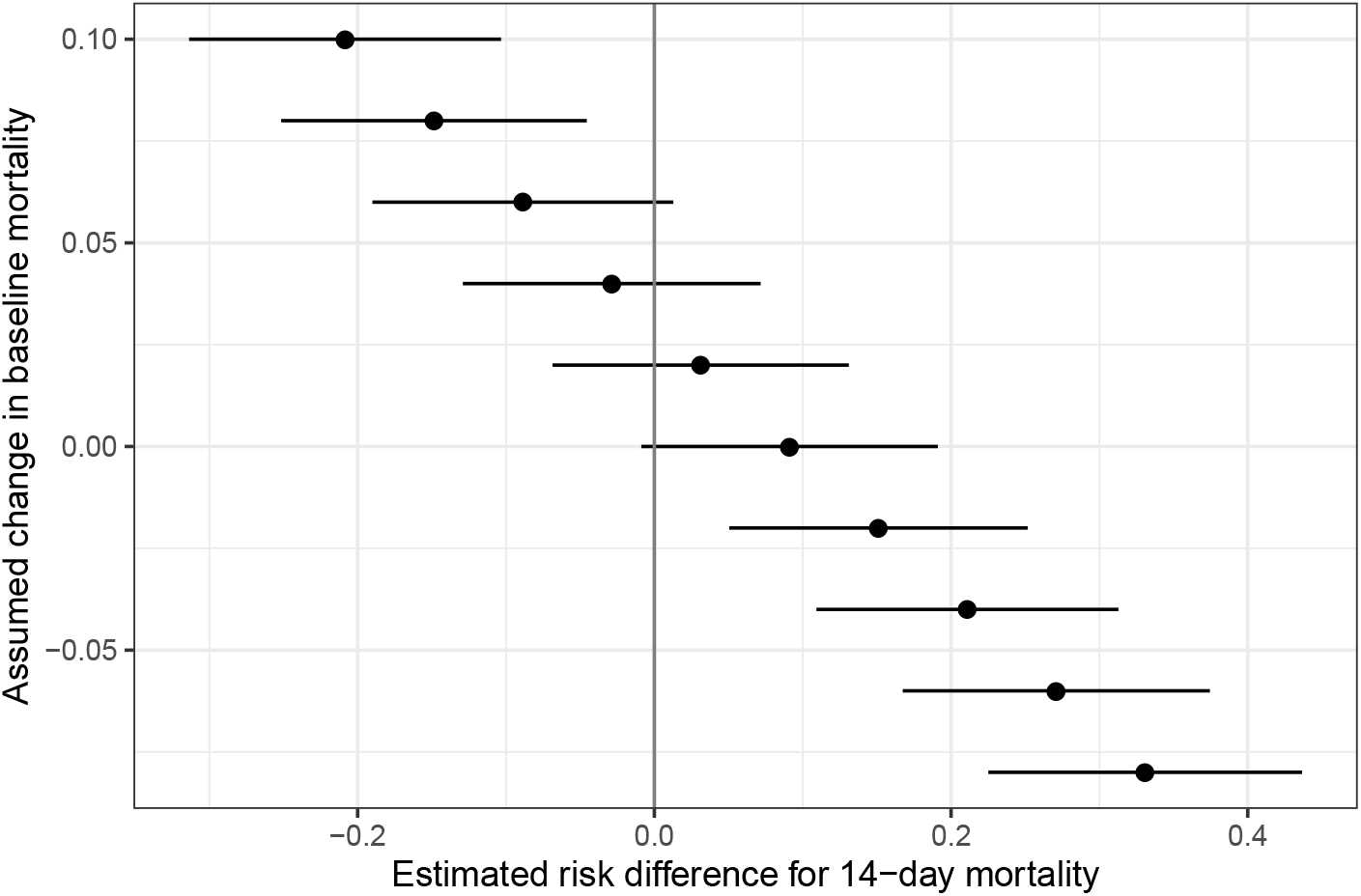
SCQE Estimates of risk difference for hydroxychloroquine. *Note:* The vertical axis indicates an assumption about the baseline trend in mortality, i.e. how mortality is postulated to have changed going from the low-use to high-use cohorts, for reasons other than changes in hydroxychloroquine use. Because the high-use cohort is the earlier one here, positive values (towards the top of the figure) correspond to falling mortality in the direction of time. At each postulated mortality trend, we see the consequent effect estimate and its 95% confidence interval.

We find that hydroxychloroquine can only be claimed to have had a significant benefit if baseline mortality decreased between the first and second cohort by 6.4 percentage points. In other words, one must argue that there was a 55% reduction in COVID-19 mortality among inpatients in this short time due to non-hydroxychloroquine related reasons, to argue for a detectable benefit of hydroxychloroquine. Second, hydroxychloroquine was significantly harmful if baseline mortality instead worsened over-time by just 0.3 percentage points—just 2.6% of the original 11.6% mortality—or more. At this boundary the point estimate for hydroxychloroquine is roughly a 10 percentage point increase in mortality. For all baseline trend assumptions in between these, we would not reject the null hypothesis of zero effect.

We also consider 28-day mortality for comparability with existing studies. For hydroxychloroquine to have been significantly beneficial would require that baseline mortality improved by 6.8 percentage points, a 47% drop from the first cohort’s 28-day mortality of 14.5%. Hydroxychloroquine would prove significantly harmful if baseline mortality rose by 0.7 percentage points.

#### Cohort comparison

In assessing the plausibility of different baseline trends it is useful to examine possible changes in the composition of the cohorts and in the treatments provided. Table 1 describes these cohorts in terms of characteristics determined prior to or very shortly after admission (A), the treatments received (B), and the predicted risk of mortality according to a range of models (C). As the purpose of such comparisons is to inform our beliefs about the plausible range of baseline mortality differences between the cohorts absent hydroxychloroquine, statistical inferences regarding the comparisons are irrelevant.

**Table 1:** Comparison of hydroxychloroquine cohorts

| A. Characteristics | Cohort means: |  |
| --- | --- | --- |
|  | High-use | Low-use |
| age (years) | 58 | 56 |
| over 65 y.o. | 36% | 33% |
| female | 42% | 46% |
| ethnicity: Hispanic | 38% | 49% |
| weight (Lb) | 194 | 187 |
| BMI (kg/m<sup>2</sup>) | 31 | 32 |
| Admit from skilled nursing facility | 4% | 9% |
| Admit from non-healthcare facility | 77% | 0.74% |
| CRP (mg/L) | 115 | 108 |
| WBC (per mcL) | 7.72 | 8.63 |
| ferritin (μg/L) | 747 | 601 |
| procalcitonin (ng/mL) | 0.89 | 1.11 |
| ICU in first 24h | 18% | 15% |

| B. Other treatments | High Use | Low Use |
| --- | --- | --- |
| remdesivir | 3% | 11% |
| tocilizumab | 7% | 3% |
| convalescent plasma | 5% | 22% |
| proning | 3% | 4% |
| dexamethasone | 4% | 5% |
| methylprednisolone | 10% | 7% |
| prednisone | 1% | 2% |
| hydrocortisone | 3% | 4% |
| nitazoxanide | 1% | 0% |

| C. Modeled risk of 14-day mortality | High Use | Low Use |
| --- | --- | --- |
| linear model (pre-tx) | 10.5% | 9.8% |
| linear model (all) | 10.9% | 9.2% |
| KRLS model (pre-tx) | 10.2% | 9.5% |
| KRLS model (all) | 10.4% | 9.1% |

(a) *Note:* Comparison of cohorts with high (earlier) or low (later) use of hydroxychloroquine, considering (A) various patient characteristics, (B) other treatments received, and (C) model-estimated risk of 14-day mortality. Lab measures (CRP, WBC, ferritin, procalcitonin) refer to the first measurement taken.

Looking first at patient characteristics prior to or shortly after admission (A), we see the two cohorts are similar overall, particularly on known risk factors such as age, gender, weight, and BMI. The proportion who identify as Hispanic rises somewhat, from 38% to 49%. Taken alone, and given documented differences in outcomes in Hispanic patients, this would contribute towards an upward shift in baseline mortality risk over time. Similarly, the fraction of patients coming from skilled nursing facilities rises somewhat (from 4% to 9%), which could also increase baseline mortality in the second cohort. Recall that, because the high use cohort precedes the low use cohort, potential increases in baseline mortality over time (as might be caused by these changes) represent negative baseline trends (i.e. moving downwards on Figure 2), and lead to more harmful estimated effects of hydroxychloroquine. On the other hand, two treatment practices (B) that could have potentially improved mortality increased over time between these cohorts: remdesivir (from 3% to 11%) and convalescent plasma (from 5% to 22%). Were these treatments to improve mortality, they would encourage us to consider possible improvements in mortality over time, moving upwards on Figure 2. Remdesivir’s effectiveness in reducing mortality remains uncertain, with the ACTT-1 trial [7] showing a benefit on time to recovery, while preliminary reports from the WHO Solidarity trial [8] show no significant mortality benefit. Nevertheless, even if these treatments are relatively effective, the change in baseline mortality due to these alone could not be large, given the low usage rates. Suppose that nearly all of the 11.6% of patients who would have died in the low-use cohort (based on the rate in the earlier, high-use cohort) received treatment with remdesivir and/or convalescent plasma. Suppose these therapies, in any combination, reduce mortality by 30%, which we take to be generous given preliminary evidence. This would produce a 3.5 percentage point drop in mortality. Assuming such a baseline trend (represented by 0.035 in Figure 2) would likely be conservative, given these assumptions and that other factors such as ethnicity suggest mortality change in the opposite direction. Yet, even at an assumed baseline trend of 0.035, hydroxychloroquine does not prove significantly beneficial and has a point estimate near zero.

Finally, these differences in the cohorts are important only insofar as they suggest different baseline mortality rates. Using simple linear probability models (C), we predict 14-day mortality using only patient characteristics prior to treatment (“Linear model (pre)”), or using those characteristics plus information on treatments (“Linear model (all)”). The same predictions can instead be made using a more flexible and powerful machine learning model, kernel-regularized least squares (KRLS, [9]). These models are reasonably predictive: the linear model with all variables explains 17% of the variation in mortality; the KRLS model with all variables explains 52%. Yet, the average modeled risk levels in the two cohorts remains similar, as shown in Table 1. The low-use (second) cohort has slightly lower risk by 0.7 to 1.7 percentage points. Such model estimates only inform the range of plausible baseline trends considered. If we consider, for example, a 1 percentage point drop in baseline mortality (a baseline trend represented by .01 on Figure 2), this corresponds to a non-significantly harmful increased risk of 6 percentage points (95% CI=[-0.04, 0.16]).

### 3.2 Dexamethasone: plausibly beneficial with very low risk of harm

In the low-use (first) cohort, 5.7% (35/614) of patients were given dexamethasone, and the 14-day mortality rate was 8.1% (50/614). In the high-use (second) cohort, 46% (287/622) of patients were given dexamethasone, and the 14-day mortality rate was 4.0% (25/622). SCQE formalizes the simple logic that while mortality fell in the higher-use cohort, this only implies a benefit of dexamethasone if mortality would not have improved too greatly “on its own.” In Figure 3, the vertical axis again represents different postulated baseline trends. Because the transition from low-use to high-use for dexamethasone is now the transition from earlier to later cohorts, positive trends indicate higher (worse) baseline mortality over time.

**Figure 3:**
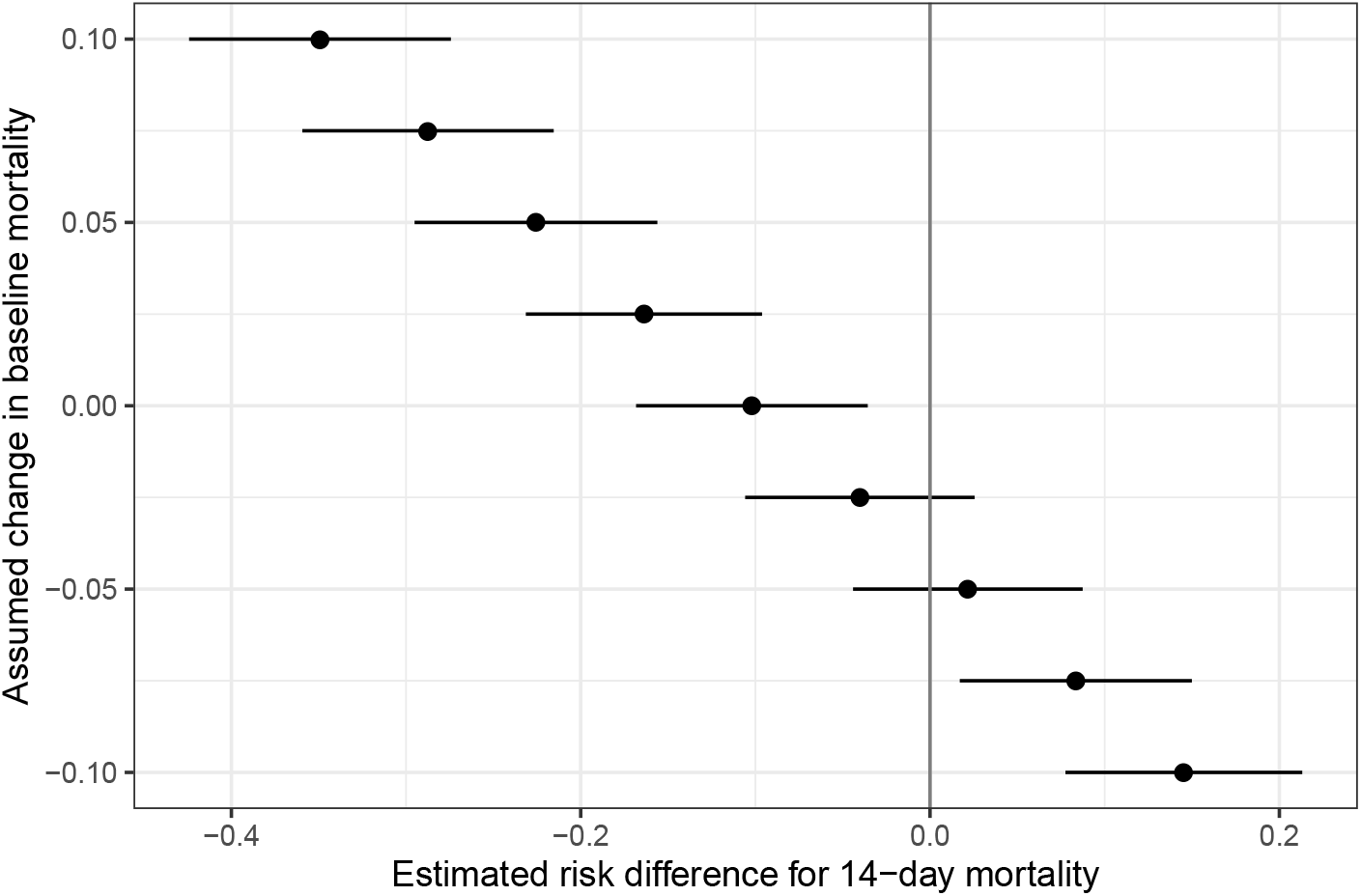
SCQE Estimates of risk difference for dexamethasone. *Note:* The vertical axis indicates an assumption about the baseline trend in mortality, i.e. how mortality is postulated to have changed going from the low-use to high-use cohorts, for reasons other than changes in dexamethasone use. Because the high-use cohort is now the later one, positive values (towards the top of the figure) correspond to increases in mortality over time. At each postulated mortality trend, we see the consequent effect estimate and its 95% confidence interval.

We find that dexamethasone had a significant benefit if baseline mortality was increasing, flat, or going down by as much as 1.5 percentage points (19%) for reasons not related to dexamethasone use. For dexamethasone to be significantly harmful, by contrast, baseline mortality had to improve by 6.8 percentage points to a mortality rate of just 1.3%, an 84% drop. As discussed below, these results support a reasonable possibility that dexamethasone had a benefit, while making it very unlikely that it was harmful.

Results are similar regarding the secondary outcome of 28-day mortality. Dexamethasone proves statistically beneficial so long as mortality rose, stayed flat, or fell by as much as 2.3 percentage points. Further, to prove harmful, baseline mortality would have to drop by 8.7 percentage points. Given the first cohort’s 28-day mortality rate of 10.9%, this would mean arguing that mortality was reduced to just 2.2% in the second cohort for reasons other than increasing dexamethasone use.

#### Cohort comparison

Table 2 aids in reasoning about possible baseline trends by comparing the high- and low-use cohorts on numerous characteristics.

**Table 2:**
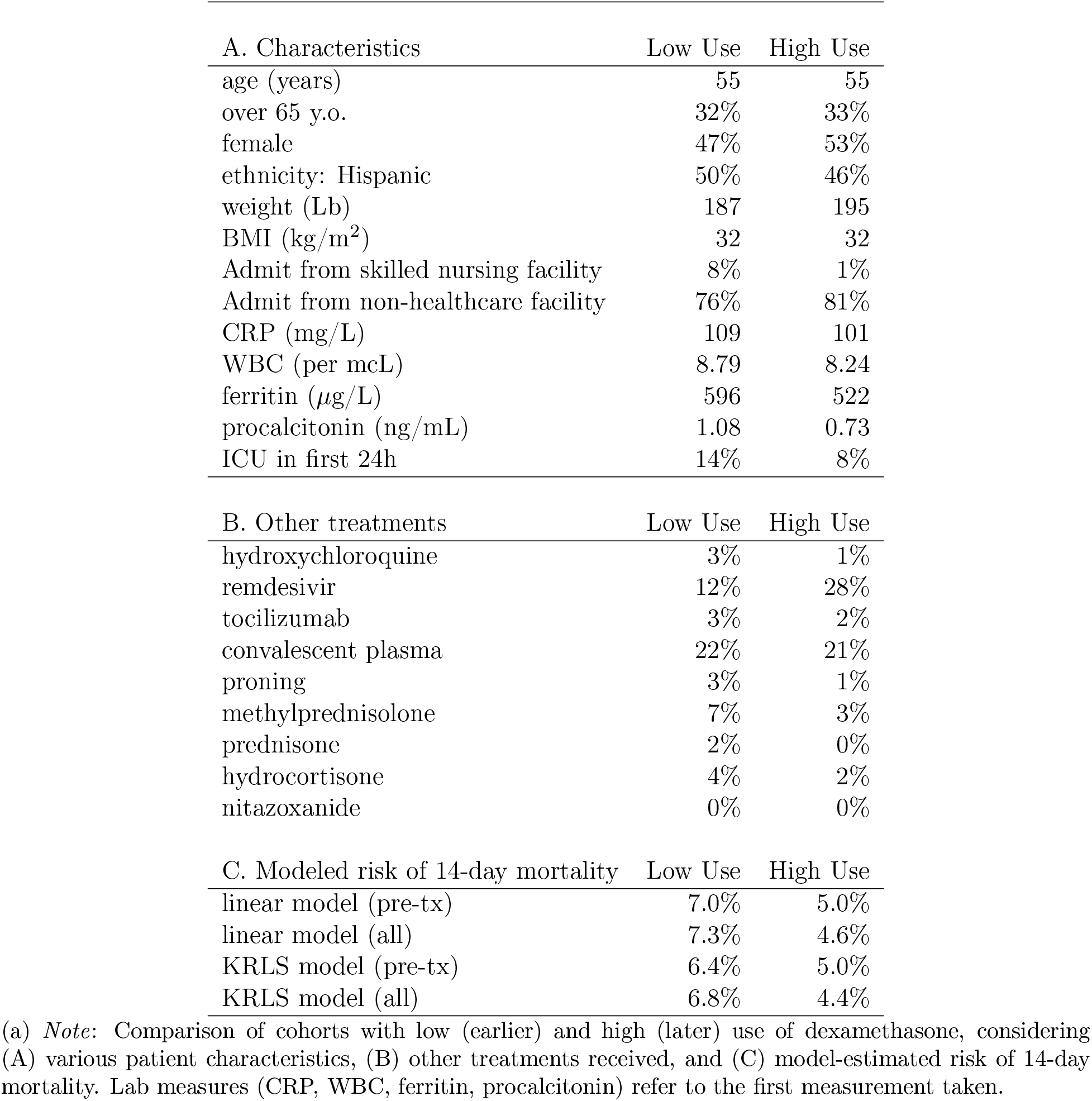
Comparison of dexamethasone cohorts

Most differences between the cohorts are small and do not revise the range of baseline trends we can consider plausible. One worrying exception is again remdesivir, with increased usage (from 12% to 28%) alongside dexamethasone. As noted above, while evidence for remdesivir’s effectiveness remains mixed [7, 8], one can conservatively examine its potential impact on baseline mortality under the assumption that it has a given benefit. If remdesivir reduced mortality by 20 percentage points, for example, the increase in usage from 12% to 28% would suggest a drop in the baseline mortality by 3.2 percentage points. If we took this to be the baseline trend (−0.032), it would suggest a benefit of dexamethasone that does not reach significance (RD = −0.023, 95% CI=[−.088, .043]).

Looking to models of mortality risk, in every case the predicted risk of mortality fell going into the second (high-use) cohort, by 1.4-2.7 percentage points. If the baseline trend was believed to be within this range, the corresponding effect estimates for dexamethasone would range from a significant beneficial RD of −0.067 to a non-significant beneficial RD of −0.035. In summary, dexamethasone could very plausibly have had either a beneficial or a null effect, while we can nearly rule out that it had a harmful one.

## 4 Discussion

Our study shows what can be inferred about the effects of hydroxychloroquine and dexamethasone use on COVID-19 mortality using only electronic health records from outside of randomized trials. We use the SCQE approach, which explicitly links assumptions about baseline trends in mortality to conclusions we can reach about a therapy’s effect on mortality.

Several considerations aid in gauging what baseline trends are (im)plausible or (im)probable, and hence the conclusions that can be supported. Studies of overall mortality in other populations show substantial decreases over time. For example [10] show large decreases in mortality in mid-April to May as compared to March among critical care patients in England, which they argue are not due to changes in patient demographics. Such results do not directly speak to the baseline trends expected in our analysis given (i) differences in the population, time period, outcomes, and (ii) that these reflect overall mortality inclusive of changes in treatments like dexamethasone, not the baseline mortality trends we require. In fact, [10] argue changes in mortality may be partly due to treatments employed in the RECOVERY trial, such as dexamethasone. Nevertheless, there are numerous reasons to expect improvements in baseline mortality in our sample due to changes in other various treatment practices over time. Though the cohorts we compared had similar exposure to most therapies (Tables 1 and 2), changes in treatment practice that remain unobserved to us could have led to improvements, notably delaying or avoiding invasive respiratory support and improved ventilator management. Given such possibilities, the reduced mortality seen in other settings, and the otherwise similar demographics and estimated mortality risk in these cohorts, we would judge small increases in baseline mortality to be unexpected but possible, while we judge large increases in baseline mortality—say by 20% or more—to be extremely unlikely.

It is more difficult to say how large a drop in baseline mortality would be too large to be plausible. For many other, longer-running diseases, it might be reasonable to suggest baseline mortality would drop by no more than perhaps 10% in a matter of months. Given COVID-19’s novelty, however, a much more generous bound is required. Still, given information about about treatment practices in this health system (two of the authors are physicians there), we do not expect any otherwise undocumented highly effective treatment was initiated and widely used in this period. While we would argue that a 40% drop in baseline mortality can neither be ruled out nor defended with certainty, we regard a drop of 75% or more to be highly improbable.

In the case of hydroxychloroquine, the mortality rate decreased as hydroxychloroquine use decreased. This alone does not imply that hydroxychloroquine was harmful; such an inference depends on how mortality would have changed anyway, i.e. the baseline trend. For example, if mortality would have fallen even faster absent the drop in hydroxychloroquine usage, then the observed data would be consistent with a beneficial effect of hydroxychloroquine. Hydroxychloroquine was detectably beneficial only if baseline mortality improved from the earlier to later cohort by 6.4 percentage points (55%). This is possible, but we must accept that it is far from confidently defensible, and certainly not supported by the modeled changes in mortality risk between these cohorts. Further, against the difficulty of defending a beneficial effect, one must consider the risk that hydroxychloroquine was harmful. If mortality worsened from one cohort to the next by even 0.3 percentage points, then hydroxychloroquine must have had a statistically significant harmful effect. These results are consistent with evidence from randomized trials testing hydroxychloroquine for early treatment of mild COVID-19 in adults [11], for reduced mortality among hospitalized patients (RECOVERY trial[12, 13]), or prophylactic protection against infection among exposed participants [14], all of which concluded hydroxychloroquine had null or potentially harmful effects on their varied outcomes.

For dexamethasone, SCQE reveals that it was significantly beneficial if baseline mortality was increasing over time between cohorts, stayed flat, or fell by up to 22% (1.5 percentage points). Baseline trends falling in this range are certainly plausible or even probable. Consequently we must conclude that dexamethasone could plausibly have had a beneficial effect in this sample. Such a potential benefit is to be weighed against the risk of harm. Here the results are clear: statistically significant evidence of harm requires that baseline mortality improved between cohorts by at least 6.8 percentage points, leaving mortality at just 1.3% in the later cohort (an 84% improvement). We regard this as highly unlikely given the small differences between the cohorts and that no undocumented but highly effective treatment was likely to have been discovered and widely used in the second cohort. Our results are consistent with, though more reserved than, conclusions drawn from the CoDEX trial [15] showing increased days alive without mechanical ventilation and the RECOVERY trial [5] showing lower mortality for those under mechanical ventilation or with oxygen supplementation at randomization. Our results, however, speak to the plausible range of effects among those given dexamethasone by choice, rather than those meeting eligibility requirements for such trials.

### 4.1 Limitations

The central limitation of this study and approach is also its strength: it avoids providing a narrow claim, because it avoids relying on a narrow assumption that is unlikely to be defensible. This may remain unsatisfying for readers accustomed to more specific claims. However, the types of estimates offered by conventional approaches obtain their apparent specificity not by providing more information, but by concealing how our estimate depend upon on the value of uncertain assumptions, thus risking over-confidence in unsupported conclusions. The SCQE approach serves to communicate what can (not) be claimed subject to what assumption, leaving the reader to argue positively for the assumptions that would be required to reach a conclusion and illustrating the limits of our knowledge.

Another limitation, specific to this study, regards sample size. While the sample is larger than those in some randomized trials, and more than sufficient for SCQE mechanically, the estimated effect has to be relatively large (roughly 10 percentage points or more) for the 95% confidence interval to exclude zero. This in turn means that our conclusions will be less decisive over a given plausible range of baseline trends than they may have been with similar estimates but a larger sample.

Finally, in both of the studies, the cohorts we defined were largely similar in their composition and use of other treatments, which is not necessary but makes it far easier to reason about plausible bounds on the baseline mortality difference between cohorts. That said, differences in the use of remdesivir remain non-trivial in both cases, with convalescent plasma use also changing in the hydroxychloroquine study. We have discussed the degree to which these could influence the baseline mortality difference, and what this means for our estimates. Still, the existence of these changes is a nuisance that widens the range of plausible estimates. A promising option suitable in some contexts for future research would be a “design-based” version of the SCQE in which hospitals plan to make a new treatment available, again by choice rather than as part of an RCT, while intentionally limiting other changes in practice or patient composition over a period of time around this transition. To the degree this is feasible, it would buoy arguments for baseline trends of smaller magnitude, resulting in a narrower range of plausible effect estimates while preserving the ability to offer patients and doctors choice in treatment.

### 4.2 Conclusions

Our results are largely consistent with those of existing trials on hydroxychloroquine and dexamethasone, despite examining outcomes for patients outside of randomized trials using only electronic health records. This study provides not only corroborative evidence from other populations regarding these treatments, but also a useful and accessible application of the SCQE approach to aid adoption.

More broadly, as observational studies are likely to remain part of the research landscape, SCQE can offer a rigorous way to understand what can (and cannot) be safely determined based on patient experiences with non-randomized treatments. SCQE estimates may be particularly useful prior to the availability of data from randomized trials, or in domains such as quality-improvement studies in which randomized trials are not always performed. This approach can also complement randomized trials, as demonstrated here, by offering corroborating evidence and assessing efficacy in a population that will often differ from those enrolled in trials.

We endorse the argument that even—or especially—in moments of urgency such as a pandemic, every effort should be made to launch and complete coordinated, well-designed randomized trials [16]. Nevertheless, there remains an important role for credible observational studies that avoid risks of producing misleadingly confident results built on fragile assumptions.

Numerous opportunities remain to apply this approach to COVID-19 therapeutics in development, including convalescent plasma, monoclonal antibodies, and additional antivirals and anti-inflammatory agents currently being used experimentally. At a time when ongoing randomized trials often coexist with parallel access to experimental therapies under expanded access provisions, the reduced uptake of randomized trials may additionally increase the importance of methodological frameworks such as SCQE to evaluate observational data.

## Data Availability

De-indentified data sufficient to replicate all analyses here will be made available at the time of publication.

## Footnotes

1 Borrowing from the potential outcomes framework [3, 4], we can conceptualize a patient’s outcome both had they taken the treatment (their treatment outcome) and had they not (their non-treatment outcome), regardless of their actual treatment status. An “average non-treatment outcome” for a cohort, then, is the average outcome we would observe had no patients received treatment.

2 This does change the interpretation of effect in terms of the population of patients to whom it applies; see [2].

3 Analyses using only aggregate data of this kind can be conducted using web-based software available at https://amiwulf.shinyapps.io/SCQE_demo/.

